# Feasibility of estimation of aortic wave intensity using non-invasive pressure recordings in the absence of flow velocity in man

**DOI:** 10.1101/2020.01.22.20018457

**Authors:** A.D. Hughes, C. Park, A. Ramakrishnan, J. Mayet, N. Chaturvedi, K.H. Parker

## Abstract

**Background:** Wave intensity analysis provides valuable information on ventriculo-arterial function, hemodynamics and energy transfer in the arterial circulation. Widespread use of wave intensity analysis is limited by the need for concurrent measurement of pressure and flow waveforms. We describe a method that can estimate wave intensity patterns using only non-invasive pressure waveforms (pWIA).

**Methods:** Radial artery pressure and left ventricular outflow tract (LVOT) flow velocity waveforms were recorded in 12 participants in the Southall and Brent Revisited (SABRE) study. Pressure waveforms were analysed using custom-written software to derive the excess pressure (*P_xs_*) which was scaled to peak LVOT velocity and used to calculate wave intensity. These data were compared with wave intensity calculated using the measured LVOT flow velocity waveform. In a separate study, repeat measures of pWIA were performed on 34 individuals who attended 2 clinic visits at an interval of approximately 1 month to assess reproducibility and reliability of the method.

**Results:** *P_xs_* waveforms were similar in shape to aortic flow velocity waveforms and the time of peak *P_xs_* and peak aortic velocity agreed closely. Wave intensity estimated using pWIA showed acceptable agreement with estimates using LVOT velocity tracings and estimates of wave intensity were similar to values reported previously in the literature. The method showed fair to good reproducibility for most parameters.

**Conclusions:** The *P_xs_* is a surrogate of LVOT flow velocity which, when appropriately scaled, allows estimation of aortic wave intensity with acceptable reproducibility. This may enable wider application of wave intensity analysis to large studies.

## Introduction

Blood pressure (BP) results almost entirely from waves generated by the heart; the intensity of these arterial waves is an important measure of ventriculo-arterial function and their interaction. While wave intensity analysis is not the only method to characterise waves in the circulation, (Westerhof et al., 2005; Caro et al., 2012) it has proved an increasingly valuable approach to understanding hemodynamics and wave propagation in the circulation, since it quantifies the intensity and energy carried by forward and backward-travelling waves, along with their timing. (Parker and Jones, 1990; MacRae et al., 1997; Parker, 2009; Broyd et al., 2015; Su et al., 2017) This has prognostic value: wave reflection has been reported to predict cardiovascular events independently of other cardiovascular risk factors (Manisty et al., 2010) and more recently, elevated wave intensity has been independently associated with greater decline in cognitive function from mid-to late life.(Chiesa et al., 2019) This latter observation is consistent with suggestions that excessive pulsatile energy transfer is responsible for microvascular damage in the cerebral circulation. (Mitchell, 2018)

Traditionally, analysis of wave intensity requires that both pressure and flow (or flow velocity) are measured, ideally simultaneously. These measurements can be onerous and technically challenging in large scale studies. We therefore examined the feasibility of deriving estimates of wave intensity based on measurement of pressure only. The method for calculating wave intensity using only pressure measurements is based on an observation made by Wang et al. (Wang et al., 2003), who reported that excess pressure (*P_xs_*), the difference between measured and reservoir pressure, was directly proportional to flow in the aortic root, *Q_in_* in dogs. Subsequent studies in humans employing invasive measurements of pressure and flow velocity in the aorta (Davies et al., 2007) and non-invasive measurements of carotid artery pressure and aortic flow (Vermeersch et al., 2009; Michail et al., 2018) have confirmed these findings. Given that wave intensity is the product of the derivatives of pressure and flow velocity this suggests that it should be possible to estimate wave intensity patterns using the measured pressure waveform and *P_xs_* derived from reservoir analysis.

We therefore examined if this approach could be used to estimate wave intensity patterns from non-invasive measurements of the pressure waveform in man using a sample from a large UK population-based longitudinal cohort, the Southall and Brent Revisited (SABRE) study. We tested whether the *P_xs_* waveform was similar to the measured aortic flow velocity waveform, assessed the agreement between wave intensity estimates made using pressure alone with the traditional approach, and also studied the test-retest reproducibility of pressure-only wave intensity analysis (pWIA) and other reservoir parameter estimates.

## Methods

Data were obtained from participants in the SABRE study, a tri-ethnic population-based cohort consisting of white European, South Asian and African Caribbean people resident in West London, UK. (Tillin et al., 2012) In brief, participants, aged 40 to 69 years, were recruited from primary care and baseline measurements performed between 1988 and 1991. Surviving participants were invited to attend a 20-year follow-up for detailed phenotyping between 2008 and 2011 and data collected at this visit were used for this study. Twelve consecutive participants who underwent measurements of aortic flow velocity by echocardiography and blood pressure waveform measurement by radial tonometry were selected to explore the feasibility of performing wave intensity analysis using a pressure-only technique (pWIA). Reproducibility of the pWIA technique was assessed on 34 participants who re-attended for study investigations within a month as part of routine quality control. Exclusion criteria for both studies were rhythm other than sinus rhythm, valvular heart disease, or any other clinical condition that prevented full participation in the study.

The study was approved by the local research ethics review committee, and all participants gave written informed consent. The study adhered to the principles of the Declaration of Helsinki and Title 45, US Code of Federal Regulations, part 46. Protection of Human Subjects. Revised November 13, 2001, effective December 13, 2001, and all procedures were performed in accordance with institutional guidelines.

### Investigations

Participants fasted and refrained from alcohol, smoking, and caffeine and were advised to avoid strenuous exercise for ≥12 hours before attendance. Participants omitted any medication on the morning of investigation. A questionnaire was completed, which detailed health behaviours, medical history, and medication. Height, weight, and waist circumference were measured as previously described. (Tillin et al., 2012) Diabetes was defined according to World Health Organization criteria, (Alberti and Zimmet, 1998) self-report of doctor-diagnosed diabetes, or receipt of anti-diabetes medication. Hypertension was defined as use of blood pressure-lowering medication from patient questionnaire and/or general practitioners’ medical record review. Coronary heart disease was defined as a coronary event or revascularization identified by medical record review and adjudicated by an independent committee. Diagnosis of stroke was based on predetermined criteria of symptoms, duration of symptoms, and MRI or computed tomography imaging from hospital admission, patient, or medical records. (Tillin et al., 2013) Heart failure, valve disease and atrial fibrillation were identified during the clinic visit and/or from medical records.

Seated brachial blood pressure was measured after 5-10 minutes rest using a validated automatic Oscillometric device (Omron 705IT). Arm circumference was measured and an appropriate sized cuff, based on British Society of Hypertension guidelines, was placed on the left upper arm. Three recordings were taken 2 minutes apart, and the second and third recordings were averaged as an estimate of clinic BP. BP waveforms were also recorded from the radial artery using a tonometer device (SphygmoCor; AtCor, Sydney, Australia) over at least six cardiac cycles, ensemble averaged and calibrated to brachial systolic and diastolic BP according to the manufacturer’s instructions. Central BP was calculated using the manufacturer’s software, which employs a generalized transfer function. Reservoir analysis (Figure 1) was performed using custom-written Matlab code (Mathworks, Inc, Natick, MA) as previously described. (Davies et al., 2014)

**Figure 1.**
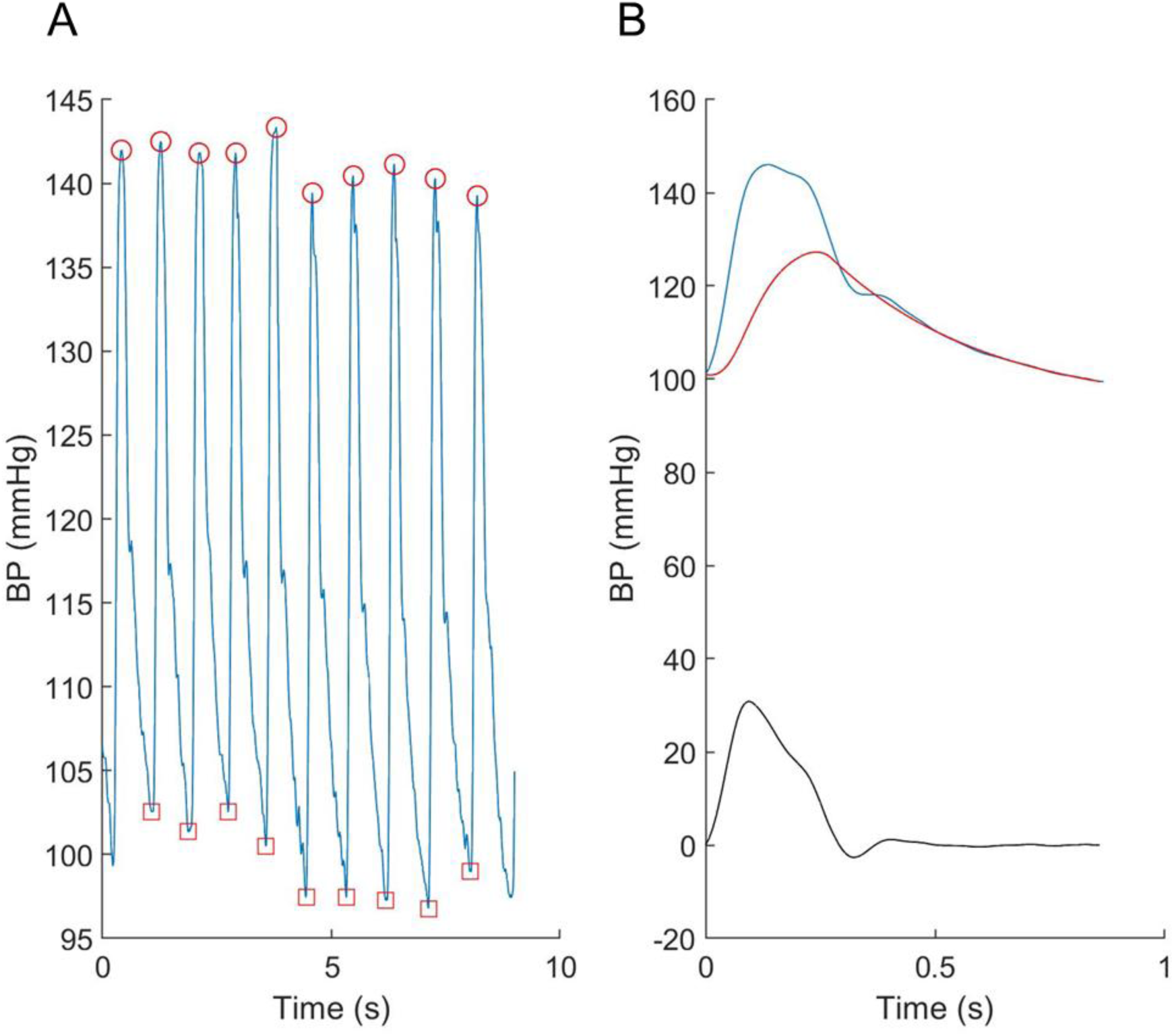
An example showing A) the individual radial pressure waveforms recorded using the tonometer and B) the ensemble averaged central pressure (blue), reservoir pressure (red) and excess pressure (black).

Echocardiography was performed using a Philips iE33 ultrasound machine (Philips, Amsterdam, The Netherlands) with a 5.0 to 1.0 phased array transducer (S5-1), as previously described. (Park et al., 2013) Aortic flow velocity was measured in the left ventricular outflow tract (LVOT) using continuous wave Doppler according to ASE/EAE guidelines (Quinones et al., 2002) and at least 3 consecutive cardiac cycles were recorded during quiet respiration. For estimation of aortic wave intensity using a conventional method, the single best quality flow velocity waveform was digitalized at a sampling rate of 200Hz and then downsampled to 128Hz to correspond to the sampling rate of the SphygmoCor data, velocity in diastole was constrained to zero. Wave intensity was calculated as the product of the derivative of the central pressure waveform and the derivative of the digitalized flow velocity waveform as previously described. (Bhuva et al., 2019)

### Fitting the reservoir and calculating pressure-only wave intensity

Details of the reservoir approach are provided elsewhere (Hughes and Parker, 2020). In brief, it is assumed that reservoir pressure, *P_res_*, satisfies overall conservation of mass for the circulation:

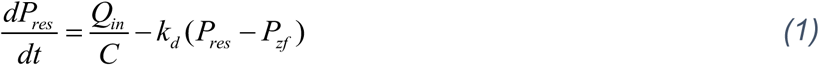

where *Q_in_* is the volumetric flow rate into the aortic root, *C* is the net compliance of the arteries, *k_d_* is the diastolic rate constant (the reciprocal of the diastolic time constant *τ = RC*, where *R* is the resistance to outflow through the microcirculation, and *P_zf_* is the pressure at which outflow through the microcirculation ceases.

The excess pressure (*P_xs_*) is the difference between the measured pressure and the reservoir pressure *(P-P_res_)* and if, as discussed above, *P_xs_* is assumed to be directly proportional to the flow in the aortic root, we can substitute

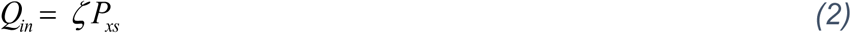

into the mass conservation equation, where *ζ* is a constant of proportionality that has some relationship with the characteristic admittance, or *1/Z_ac_*, (i.e. the inverse of the characteristic impedance) and has units of admittance.

If we define *k_s_ = ζ/C* and *k_d_ = 1/RC*, then equation (1) can be written as:

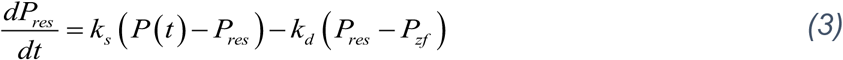

*P_res_* is given by a first-order linear differential equation:

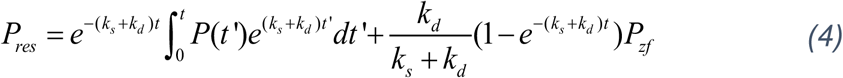

which is solved in two steps 1) by fitting an exponential curve to the pressure during diastole to estimate the diastolic parameters *k_d_* and *P_zf_*, assuming aortic inflow is zero:

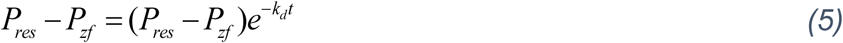

*k_s_* is then estimated by minimising the squared error between *P* and *P_res_* obtained over diastole.

Wave intensity (*dI*) is the total rate of working, i.e. the power, per unit cross-sectional area of an artery due to the pressure, *P*, with the blood flowing with velocity *U*. If flow velocity, 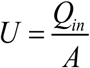 where *A* is the cross-sectional area of the aorta, and *ζ· A = ρ · c*, where *ρ* is the density of blood (assumed to be 1060 kg.m^-3^) and *c* is the wave speed. Then

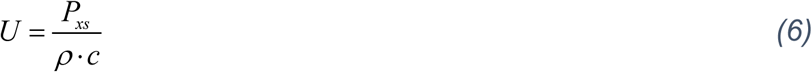

and

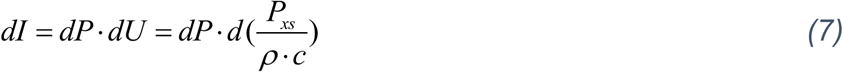

If flow velocity is measured then *c* can be estimated from equation (6). In addition to wave intensity, wave reflection index (WRI: the ratio of the area of the reflected wave to the early systolic incident wave), the ratio of peak forward to peak backward pressure (*P_b_/P_f_*) and the ratio of backward to total pressure, termed reflection index (RI) were also calculated. (Westerhof et al., 2005)

For this analysis we chose *a priori* to use *P_xs_* calculated from the estimated aortic (central) pressure waveform, as it was assumed to correspond more closely to aortic excess pressure. For the purposes of direct comparison with the conventional method *P_xs_* was calibrated to the peak aortic flow in each individual; however we also investigated the agreement when peak aortic velocity was assumed to be 1m/s in all cases. All analyses were performed using custom written software in Matlab (R2019a, The Mathworks Inc.)

### Repeatability and reproducibility of blood pressure

Reproducibility (test-retest) data for reservoir pressure, *P_xs_* and estimated wave intensity was performed on 34 participants (age 69.8 (SD =5.6) years; 26 male) who attended on two occasions separated by an interval of approximately a month.

### Statistical analysis

Statistical analyses were performed using Stata 15.1 (StataCorp, College Station, Texas, USA). Continuous variables derived from the samples were summarised as mean±SD. Reproducibility data were analysed using Bland–Altman analysis and presented as mean differences with limits of agreement (LOA). (Bland and Altman, 2003) Concordance or reliability was summarised using Lin’s concordance coefficient (rho) (Steichen and Cox, 2002) and was classified as: <0.40 - poor; between 0.40 and <0.59 - fair; between 0.60 and 0.74 - good, and >0.75 - excellent. (Cicchetti et al., 2006)

## Results

### Comparison of aortic velocity waveform with *P_xs_*

The characteristics of the 12 participants in this study are shown in Table 1. Reservoir analysis was successful in all cases and the fits were excellent (r^2^ for exponential fit in diastole = .98 (SD 0.02)). The *P_xs_* waveforms agreed fairly closely with the LVOT flow velocity waveforms measured using ultrasound (Figure 2 and Supplementary Figure S1) and there was evident correlation between the systolic upstroke of both waveforms. This was confirmed by the close agreement between the time of peak of *P_xs_* and time of peak aortic flow velocity (mean difference = 0.00 (LOA −0.02, 0.02)s). Wave intensity estimated using *P_xs_* gave the typical pattern consisting of a large forward compression wave (Wf1) in early systole, followed by a small backward wave (Wb1 - reflected wave), and followed by a moderate sized forward decompression wave in protodiastole (Wf2). Bland Altman plots indicating the agreement between peak wave intensities for conventional WIA and pWIA are shown in Figure 3 (mean difference Wf1 = −15 (LOA −106, 75) W/m^2^ × 10^4^/cycle^2^, rho = 0.83; Wb = −49(LOA −29, 19) W/m^2^ × 10^4^/cycle^2^, rho = 0.42; −44 (LOA −13, 44) W/m^2^ × 10^4^/cycle^2^, rho = 0.73). When a constant peak aortic velocity of 1m/s was assumed, the agreement was similar or only marginally worse (Supplementary Figure S2).

**Table 1.** Characteristic of participants in the study comparing wave intensity estimated using aortic flow velocity waveforms compared with excess pressure (*P_xs_*) (N = 12).

| Variable | Mean / N (SD)/[%] |
| --- | --- |
| Age, y | 65.7 (6.0) |
| Male sex, N [%] | 12 [100] |
| Ethnicity, N [%] |  |
| European | 4 [33] |
| South Asian | 8 [66] |
| Height, cm | 169.1 (7.4) |
| Weight, kg | 76.8 (13.1) |
| Diabetes, N [%] | 5 [42] |
| Hypertension, N [%] | 10 [83] |
| Systolic BP, mmHg | 147.8 (14.1) |
| Diastolic BP, mmHg | 88.3 (10.9) |
| Heart rate, min<sup>-1</sup> | 64.4 (10.8) |
| $V_{max}$, cm.s<sup>-1</sup> | 137.8 (16.9) |
| Time ($V_{max}$), s | 0.09 (0.01) |
| $P_{xs}$, mmHg | 40.1 (8.5) |
| Time ($P_{xs}$), s | 0.09 (0.01) |

Abbreviations: BP, blood pressure; $P_{xs}$ , excess pressure; $V_{max}$ , maximum aortic velocity.

**Figure 2.**
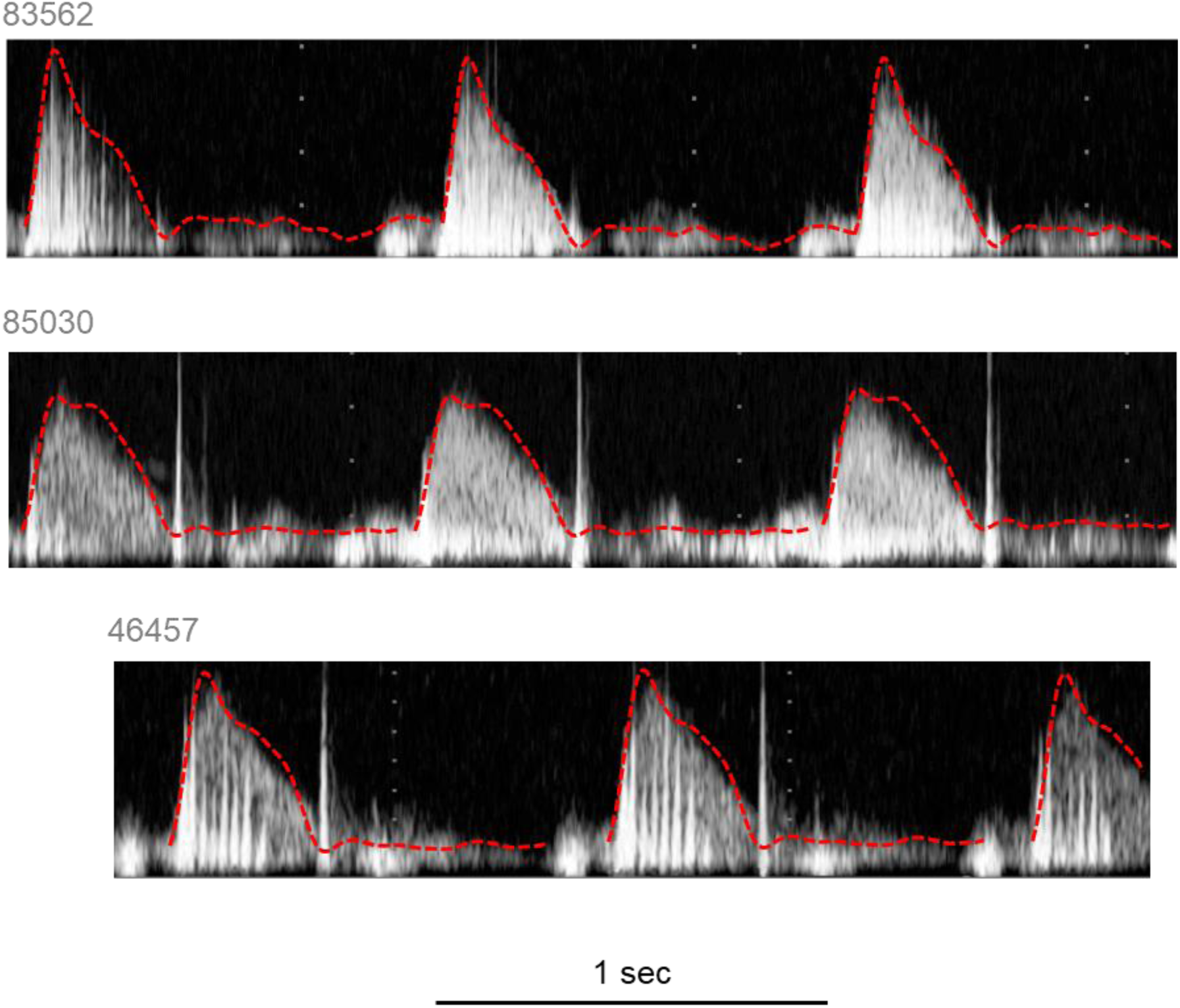
Example traces of velocity measured in the left ventricular outflow tract with the respective *P_xs_* superimposed. *P_xs_* waveforms were scaled to correspond with the peak of the aortic flow waveform.

**Figure 3.**
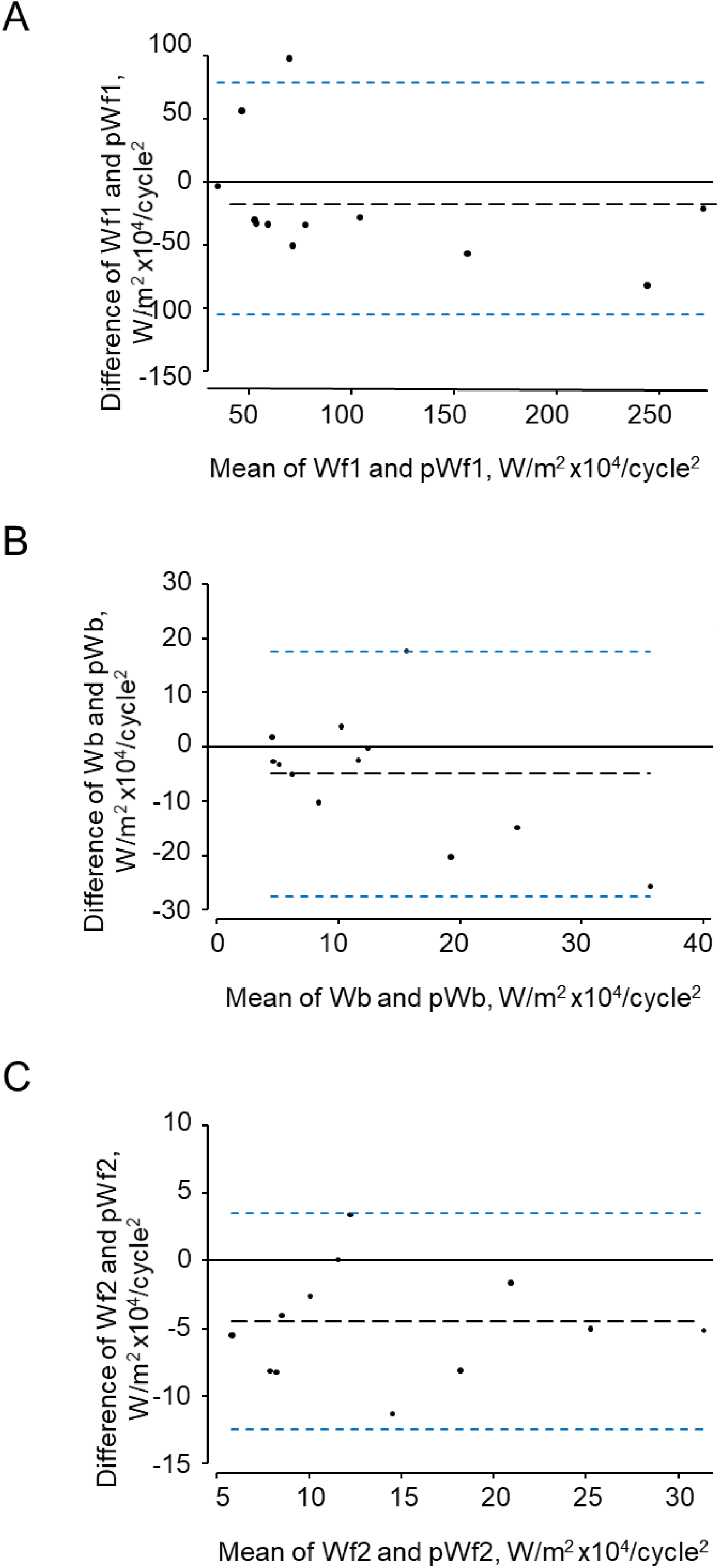
Bland-Altman plots showing agreement between the three major waves identified by traditional (Wf1, Wb, Wf2) and pressure-only wave intensity analysis (pWfl, pWb, pWf2).

### Reproducibility study

The characteristics of the 34 participants in the reproducibility study are shown in Table 2. Reservoir fitting and wave intensity calculation failed quality control in 3 and 5 cases at visit 1 and 2 respectively (12% failure rate), largely due to poor quality tonometry traces. An example of test-retest recordings in a single individual (selected to have a difference in the forward compression wave similar to the average difference) is shown in Figure 4. Bland Altman plots of all intra-individual differences for the three major waves, Wf1, Wb, Wf2 are shown in figure 5). The reliability of wave intensity was good, except for Wf2 which was poor and showed evidence of correlation between the difference and the mean (r = 0.79). Results for other measures are shown in Table 3; most showed fair or good reliability.

**Table 2.**
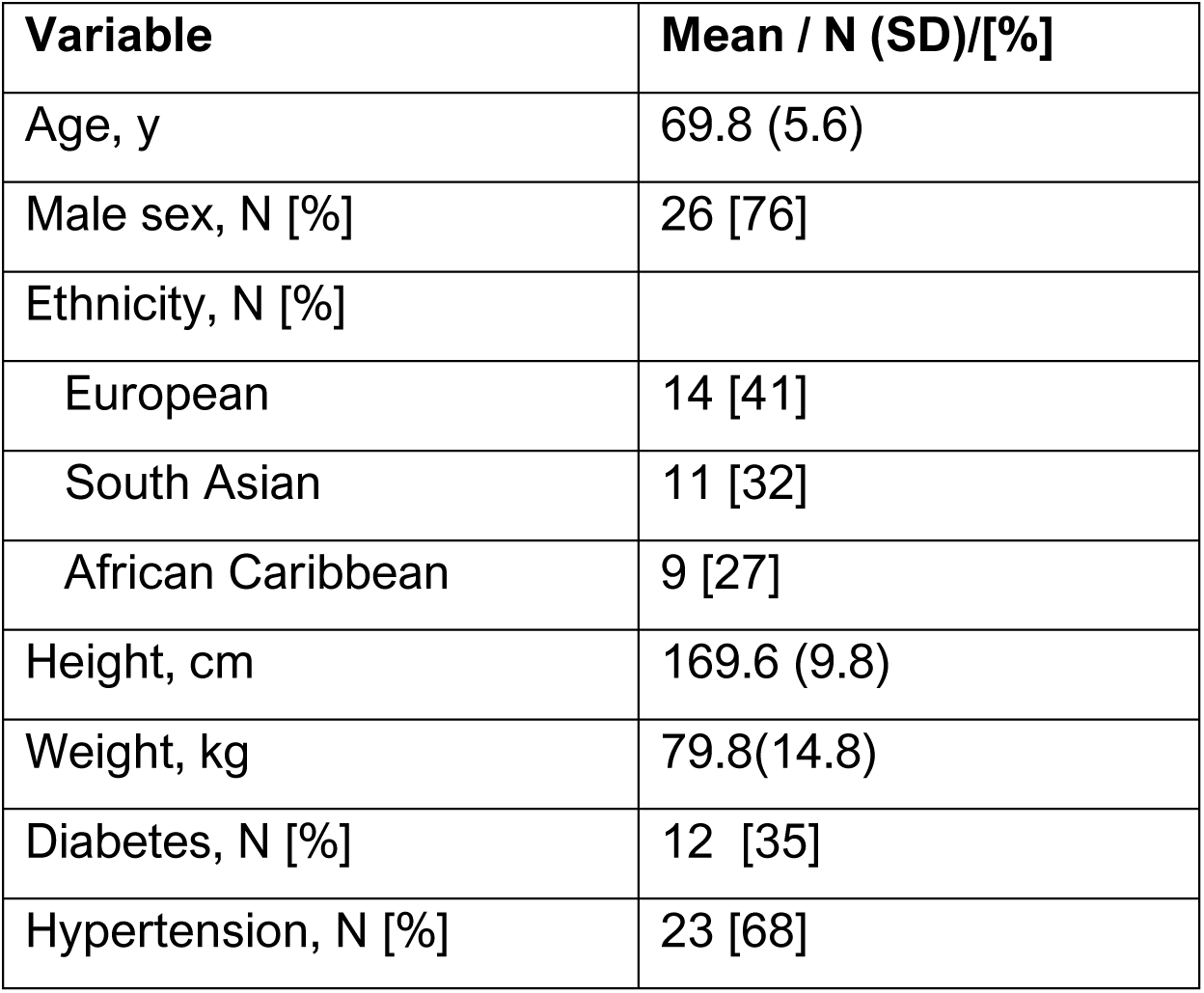
Characteristic of participants in the reproductivity study (N = 34).

**Figure 4.**
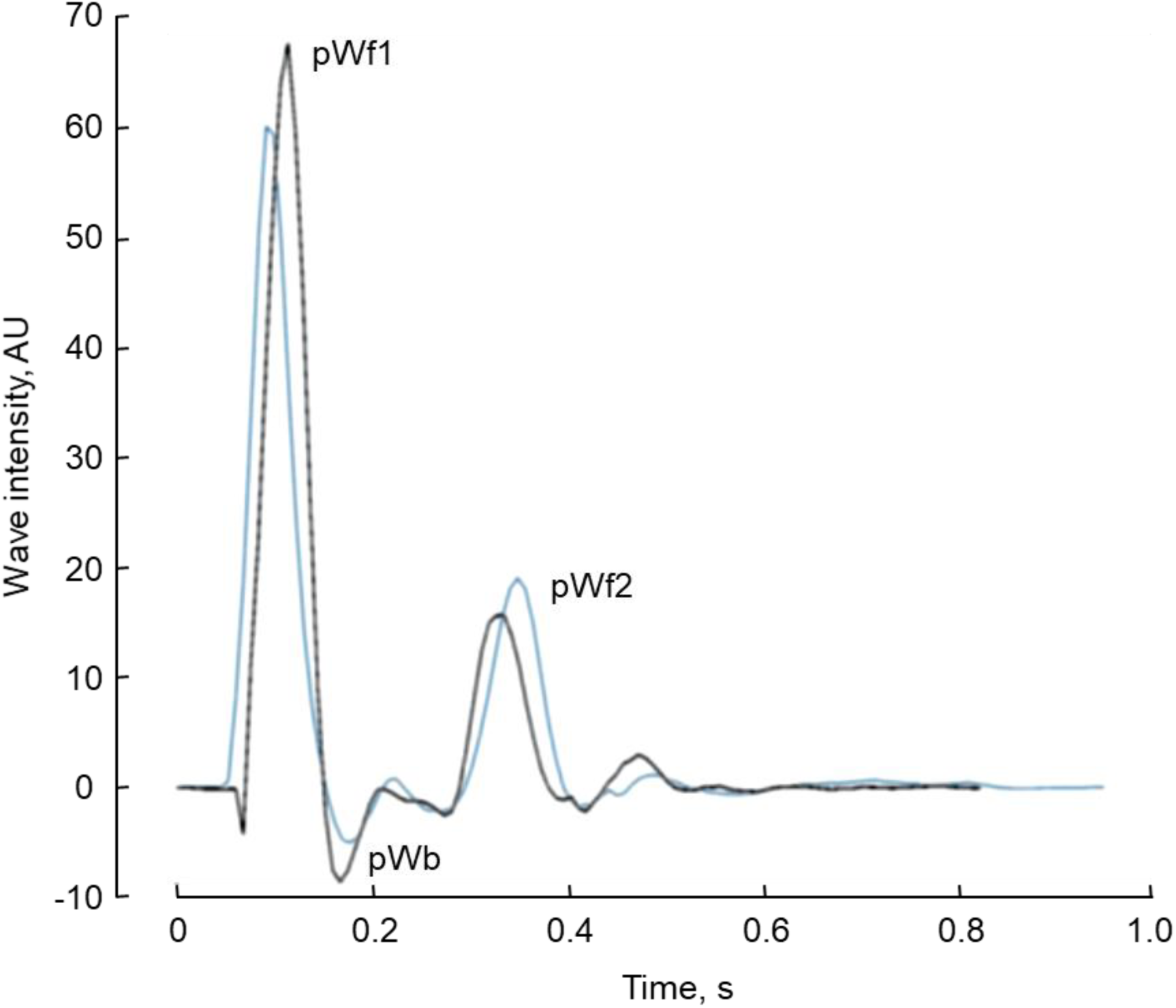
An example of pressure-only wave intensity traces recorded from the same individual on two occasions approximately 1 month apart (first visit (black) and second (repeat) visit (blue) traces). The three major waves, pWf1, pWb and pWf2 are indicated.

**Figure 5.**
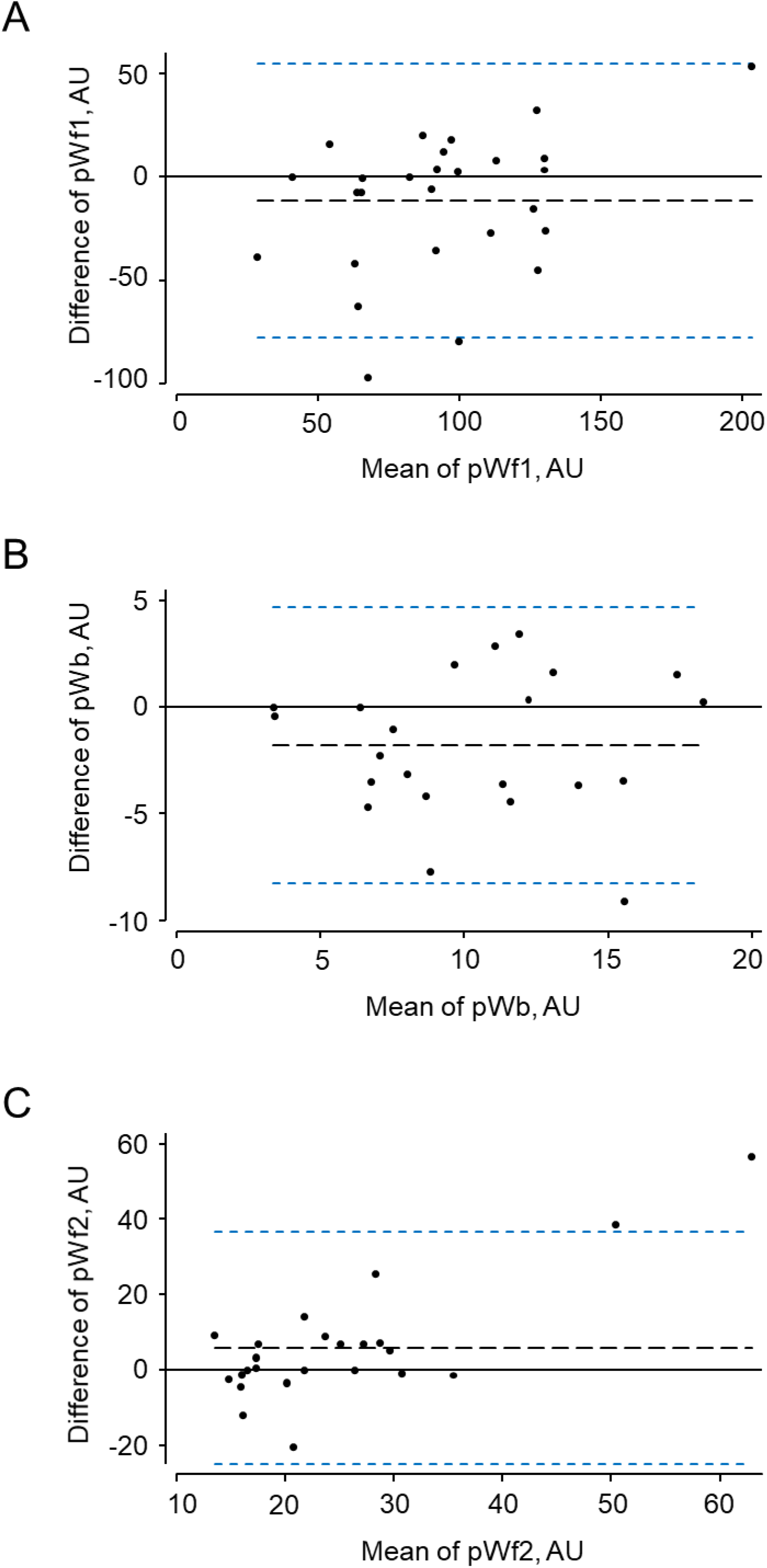
Bland-Altman plots for test-retest data of the three major waves identified by pressure-only wave intensity analysis (pWf1, pWb, pWf2) calibrated to peak aortic velocity in each individual.

**Table 3.** Results and reproductivity of key parameters measured at 2 visits.

| Variables | Visit 1 |  |  | Visit 2 |  |  | Reproducibility |  |  |  |  |
| --- | --- | --- | --- | --- | --- | --- | --- | --- | --- | --- | --- |
|  | N | mean | SD | N | mean | SD | mean difference | LOA | LOA | rho | Reliability grading |
| Systolic BP, mmHg | 34 | 136.6 | 10.1 | 34 | 134.4 | 13.8 | 2.1 | -23.5 | 27.7 | 0.42 | Fair |
| Diastolic BP, mmHg | 34 | 82.7 | 10.0 | 34 | 80.8 | 8.7 | 1.9 | -13.5 | 17.2 | 0.64 | Good |
| Heart rate, bpm | 34 | 63.4 | 9.6 | 34 | 61.1 | 9.9 | 2.3 | -10.6 | 15.2 | 0.75 | Excellent |
| maximum $P_{res}$, mmHg | 31 | 113.1 | 8.7 | 29 | 111.7 | 11.8 | 0.9 | -19.4 | 21.2 | 0.48 | Fair |
| maximum $P_{xs}$, mmHg | 31 | 36.4 | 9.1 | 29 | 36.1 | 10.1 | 0.3 | -13.8 | 14.5 | 0.71 | Good |
| Time maximum $P_{xs}$, s | 31 | 0.10 | 0.01 | 29 | 0.10 | 0.01 | 0.00 | -0.03 | 0.03 | 0.24 | Poor |
| Time maximum BP, s | 31 | 0.14 | 0.04 | 34 | 0.14 | 0.04 | 0.00 | -0.09 | 0.09 | 0.39 | Poor |
| $k_s$, s<sup>-1</sup> | 31 | 7.90 | 2.31 | 29 | 7.65 | 1.78 | 0.01 | -3.73 | 3.80 | 0.55 | Fair |
| $k_d$, s<sup>-1</sup> | 31 | 2.68 | 1.08 | 29 | 3.11 | 2.87 | 0.24 | -1.55 | 2.02 | 0.54 | Fair |
| $P_{zf}$, mmHg | 31 | 74.1 | 11.3 | 29 | 69.3 | 20.4 | 1.13 | -16.8 | 19.1 | 0.66 | Good |
| Wf1, AU | 31 | 90 | 47 | 29 | 100 | 38 | -11.6 | -77.7 | 54.5 | 0.61 | Good |
| Wb, AU | 31 | 10 | 6 | 29 | 12 | 5 | -1.8 | -8.2 | 4.7 | 0.68 | Good |
| Wf2, AU | 31 | 29 | 19 | 29 | 21 | 8 | 6 | -24.8 | 36.7 | 0.34 | Poor |
| WRI | 31 | 0.14 | 0.11 | 29 | 0.12 | 0.04 | 0.02 | -0.17 | 0.21 | 0.44 | Fair |
| $P_b/P_f$ | 31 | 0.72 | 0.04 | 29 | 0.71 | 0.04 | 0.00 | -0.08 | 0.08 | 0.53 | Fair |
| RI | 31 | 0.42 | 0.01 | 29 | 0.42 | 0.01 | 0.00 | -0.03 | 0.03 | 0.52 | Fair |

Abbreviations: AU, arbitrary units; BP, blood pressure; $k_d$ , diastolic rate constant; $k_s$ , systolic rate constant; LOA, 95% limits of agreement; $P_b/P_f$ , ratio of backward to forward pressure; $P_{res}$ , reservoir pressure; $P_{xs}$ , excess pressure; $P_{zf}$ , estimated zero-flow pressure; rho, Lin's concordance coefficient; RI, reflection index; $V_{max}$ , maximum aortic velocity; Wb, backward compression wave; Wf1, forward compression wave; Wf2, forward decompression wave; WRI, wave reflection index.

## Data Availability

The data that support the findings of this study are available from the corresponding author, [ADH], upon reasonable request.

## Discussion

We found that the *P_xs_* waveform is an acceptable surrogate of LVOT (aortic) flow velocity waveform, and, following calibration of the *P_xs_* waveform, it was possible to calculate wave intensity patterns from recordings of the pressure waveform made at the radial artery without measurements of flow. Wave intensity and related parameters estimated in this way showed acceptable agreement with conventionally measured wave intensities and the reproducibility and reliability of pWIA was similar to or better than the reliability of systolic BP. The only exception was Wf2 which showed poor reliability; this may relate to the small size of this wave and the variability in duration of ejection which introduces noise into the ensemble average of the waveform in late systole. Improved methods of ensemble averaging might be useful if Wf2 were a parameter of particular interest in a given study.

The method used to derive the flow velocity waveform from the measured pressure has some similarities with the approach used to derive flow waveforms in the ARCSolver method, which is based on a 3-element Windkesel model plus a minimal work criterion. (Hametner et al., 2013) The ARCSolver method has been reported to outperform a simple triangular flow assumption in terms of pressure separation, (Hametner et al., 2013) and it has been used for wave intensity analysis in one study, (Hametner et al., 2017) although we are not aware of any validation studies using this approach.

In our studies the observed wave intensity patterns using the pressure-only approach were very similar to those reported previously using invasive or non-invasive methods based on measurement of pressure (or diameter) and flow velocity. (Parker and Jones, 1990; Koh et al., 1998; Niki et al., 1999; Zambanini et al., 2005; Bhuva et al., 2019) The major disadvantage of the current approach is the lack of absolute calibration in the absence of a flow velocity measure. In many studies peak aortic or LVOT peak flow velocity may be measured as part of the echocardiography protocol, and this can be used to calibrate *P_xs_*. When the data were calibrated in this way the intensity of the waves agreed with those calculated using conventional methods and the values were similar to those previously reported in the literature allowing for differences in form of units. (Koh et al., 1998; Niki et al., 1999; Zambanini et al., 2005; Bhuva et al., 2019) Thus it may not be necessary to record the entire aortic velocity waveform, the peak velocity appears sufficient. However, if there is no measure of aortic flow velocity the issue of calibration is more problematic. One possibility may be to use an assumed aortic flow velocity based on previous studies. Dalen et al. (Dalen et al., 2010) measured LVOT peak velocity in 1266 healthy participants in the Nord-Trøndelag Health (HUNT) study (663 female, age range <30 to >70 years); they found no convincing evidence of a relationship between age and peak LVOT velocity in either men or women. The average LVOT velocities in this study were 98(SD 18)cm.s^-1^ in men and 101 (SD 16)cm.s^-1^ in women. Choi et al. (Choi et al., 2016) reported findings from 1003 healthy Korean adults (age 20-79y). LVOT peak velocities were slightly higher in women (men = 96(SD 15) cm.s-1 vs. women = 99(SD 16) cm.s-1) and there was a small positive relationship between increased age and higher peak LVOT velocity (corresponding to ~4 cm.s^-1^ and ~10cm.s^-1^ difference in peak velocity between age 21 to 30 and 71 to 80 in men and women respectively). Previous smaller studies have reported that peak flow velocity in the LVOT shows little or no association with body size, sex, blood pressure, or body mass index. (Gardin et al., 1987; van Dam et al., 1987; Swinne et al., 1996) This suggests that it might be possible, at least in individuals without established cardiac disease, to use an assumed peak aortic velocity to calibrate the *P_xs_* waveform. Based particularly on the more recent large population studies in Norway and Korea, (Dalen et al., 2010; Choi et al., 2016) ~100cms^-1^ seems a reasonable estimate for an assumed peak velocity in the LVOT, although a more detailed systematic review and meta-analysis on this question would be valuable.

Our study has several other limitations. It employs a small sample based on existing data using tonometry to record the BP waveform and the approach requires further validation if it is to be used in future studies employing other methods to measure pressure waveforms. We have found that the method can also be applied to pressure waveforms captured using a cuff-based arm BP monitor (unpublished data) which could allow for more automated approaches and even the possibility of measurement of ambulatory 24-hour wave intensity. The method could also be applied to invasive data where only pressure has been measured, for example in investigations of suspected pulmonary hypertension. While outside the scope of this article, it is noteworthy that a related approach could be used to estimate wave intensity in circumstances where only flow velocity was measured if accompanied by measurements of clinic BP.

In conclusion, use of the *P_xs_* waveform as a surrogate of LVOT flow velocity, when appropriately scaled, permits estimation of aortic wave intensity based on non-invasive measurement of pressure waveforms. This technique shows acceptable reproducibility and should allow wider application of wave intensity analysis to large scale trials and observational studies.

## Acknowledgements and funding sources

The SABRE study was funded at baseline by the UK Medical Research Council (MRC), Diabetes UK, and the British Heart Foundation (BHF). The follow-up study in 2008 and 2011 was funded by the Wellcome Trust (067100, 37055891 & 086676/7/08/Z), the BHF (PG/06/145, PG/08/103/26133, PG/12/29/29497 & CS/13/1/30327) and Diabetes UK (13/0004774).

AH receives support from the British Heart Foundation, the Economic and Social Research Council (ESRC), the National Institute on Aging, the National Institute for Health Research University College London Hospitals Biomedical Research Centre, the UK Medical Research Council and works in a unit that receives support from the Medical Research Council. CP is supported by a grant from the BHF. AR is supported by a grant from the BHF.

